# Telehealth usage patterns among pediatric neurology patients during and after the COVID-19 pandemic in Washington state

**DOI:** 10.1101/2025.06.04.25329018

**Authors:** Peter Y. Ch’en, Dwight Barry, Lindsey A. Morgan, Mark S. Wainwright, Payal B. Patel

## Abstract

**Background:** The COVID-19 pandemic accelerated the adoption of telehealth across the United States. While telehealth usage during the pandemic has been described, evaluating utilization across different policy periods is necessary to understand which groups engage most with this care modality under select situational barriers. This study aimed to analyze telehealth usage by demographic groups in a pediatric neurology cohort, focusing on utilization patterns during and after the pandemic.

**Methods:** A retrospective analysis was conducted of 35,700 visits among 18,053 unique pediatric neurology patients at Seattle Children’s Hospital across three phases of the pandemic (initial – 3/2020-5/2020, sustainment – 6/2020-3/2022, and maintenance – 4/2022-3/2023). Logistic regression models assessed the impact of demographic factors on telehealth utilization. Regression effect sizes were estimated for language, race/ethnicity, and insurance type.

**Results:** Telehealth utilization varied across demographic groups and geographic regions. Non-English speakers had lower telehealth usage during the sustainment and maintenance phases (OR 0.56; OR 0.50, respectively). Black patients had lower usage in the sustainment phase (OR 0.86). Patients with government insurance had decreased usage during all phases compared to those with commercial insurance (OR 0.91; OR 0.83). Patients residing farther from urban centers consistently had higher telehealth utilization across all phases.

**Conclusions:** This study identified differences in telehealth utilization by language, race/ethnicity, insurance, and geographic location. The cohort reflects the demographic makeup of Washington state, including significant rural representation. Telehealth remains especially important for rural communities, highlighting the need to preserve and enhance access - particularly through language support - to ensure equitable care delivery statewide.

## Introduction

Telehealth has the potential to improve access to neurological care for children, especially during a pandemic when in-person visits may be limited. The ability to safely provide remote telehealth care to patients during the COVID-19 pandemic helped maintain continuity of care for children.^1^ For families in rural areas that have limited access to care, telehealth eliminates the need for long-distance travel and time away from school for children and work for parents. The rapid expansion of telehealth during the COVID-19 pandemic helped continue care, particularly for vulnerable populations such as children living with disability, during a public health crisis.

Despite its advantages, there are significant barriers that may prevent equitable access to telehealth across different populations. Investigating telehealth utilization patterns can help identify interventions to promote equitable access.^2^ Proactively identifying telehealth barriers can also help anticipate what populations are most at-risk of care gaps during future public health emergencies.^2^ Understanding these differences is essential to ensuring equitable healthcare delivery and improving health outcomes.^3^ One of the most common challenges identified in published literature is barriers to technological access including the lack of high speed broadband internet and supported devices that are optimal for telehealth visits.^5^ Children from lower-income families are less likely to use telehealth services, and language barriers can further restrict access for non-English-speaking families. Telehealth has the potential to bridge some of the common gaps in care, but factors such as resource limitations, digital literacy, language barriers, cultural preferences, and socioeconomic factors contribute to disparate and inequitable access and/or utilization of telehealth. Keeping these barriers in mind when analyzing general telehealth utilization patterns is crucial for contextualizing these results and helping create targeted interventions for the most vulnerable in our population.^6^

Washington state’s rural communities in particular face unique healthcare challenges that make telehealth particularly valuable. Rural Washington comprises approximately 16% of the state’s population but covers over 80% of its landmass, creating significant geographic barriers to specialty care access.^7–9^ These vast distances mean that many residents must travel long hours to reach healthcare facilities, often in difficult weather conditions, which can delay or deter necessary care. Washington state has also recognized primary care physician shortages in rural areas as a significant public health concern, with projections indicating a 7% decrease in rural primary care physicians per capita by 2025.^7^ Telehealth in particular is an appealing solution that may be able to help bridge these provider gaps by facilitating real-time specialist consultations for patients without increased travel burden. These demographic and geographic considerations make Washington state an ideal setting to examine telehealth utilization patterns and their implications for healthcare delivery.

In this study, telehealth usage patterns were analyzed across the pandemic and post-pandemic in the division of pediatric neurology at Seattle Children’s Hospital in Seattle, WA. The study aimed to systematically analyze usage of telehealth by demographic groups across various time periods during the pandemic to identify differences in telehealth usage among the population served. The patterns of telehealth use observed in this study underscore telehealth’s critical role in supporting communities across Washington State and emphasize the ongoing need to maintain accessible telehealth services for these populations.

## Methods

### Study Design and Patient Eligibility

A retrospective analysis was conducted of 35,700 visits among 18,053 unique patients in the pediatric neurology division at Seattle Children’s Hospital in Seattle, WA from the onset of the COVID-19 pandemic in March 2020 to March 2023. Patients up to the age of 21 years old were eligible for study inclusion. Data was extracted via a query of an institutional instance of Tableau, a data management and analytics software used in part for quality improvement at Seattle Children’s Hospital. This study was considered to be exempt by the Seattle Children’s institutional review board given that the goal of this study is for quality improvement in care delivery and that the patient data used were deidentified at onset.

The study time period was split into three pandemic related phases: initial – March 2020-May 2020, sustainment – June 2020-March 2022, and maintenance – April 2022-March 2023. These phases were determined based on the quality improvements efforts to access telemedicine that were implemented at Seattle Children’s Hospital throughout the pandemic related to milestone events such as lock down restrictions (initially developed in March 2020 and with sustained policies in place beginning in June 2020) and statewide access to pediatric formulations of the COVID-19 vaccine (starting in April 2022). Clinical demographics (age, sex, self-assigned race/ethnicity, language, location), medical complexity categorization, insurance status, and appointment types (in-person, telehealth, telephone) were extracted, across all appointments by pandemic phase. Patients were classified by medical complexity using the Pediatric Medical Complexity Algorithm (PMCA), which categorizes conditions as Complex Chronic (multisystem disorders), Noncomplex Chronic (single-system conditions), No Chronic Disease, or Not Categorized.^10^ Geographic home locations were defined via the Rural-Urban Commuting Area (RUCA) framework, distinguishing urban cores (e.g., Seattle/Tacoma) from rural regions (e.g., “ rural” for areas >90 minutes from Seattle or Tacoma).^11^

### Statistical Analysis

Descriptive statistics are presented as median (interquartile range [IQR]) and n (percent). Mixed effects logistic regression was used to assess the probability of having a telehealth appointment for each of the three time periods by race and ethnicity, language group, insurance type, home location, and patient complexity (PMCA category), accounting for multiple visits by the same patient via random intercepts. Regression effect sizes were then estimated across several significant factors (language, race/ethnicity, insurance type) in relation to telehealth utilization. Analysis was performed using R version 4.4 (R Core Team 2024).

## Results

This study analyzed 35,700 pediatric neurology visits at Seattle Children’s Hospital across three pandemic phases: initial (n=2,956), sustainment (n=24,498), and maintenance (n=8,246), with a total of 18,053 unique patients (**Table 1**). The patient population had a median age of 10 years (IQR: 5-15) across all phases, with a consistent gender distribution of approximately 52% female and 48% male patients. Non-Hispanic White patients comprised of the largest group across all phases of the pandemic (~54%). Of all patient visits (both telehealth and in-person), 34% had complex chronic conditions. More patients with complex chronic conditions were seen during the initial phase (40%) compared to the maintenance phase (28%). Commercial insurance was the predominant coverage type (56% overall), with government insurance accounting for 44% of patients. Most patients (91%) were predominantly English speakers. Geographically, the majority of patients (60%) were from the Seattle/Tacoma/Everett area. In-person appointments were lowest in the initial phase (26%) and steadily rose to 48% in sustainment and 64% in the maintenance phase.

**Table 1.** Demographics by each patient visit.

| **Characteristic** | **Overall**<br>**N = 35,700<sup>I</sup>** | **Initial**<br>**N = 2,956<sup>I</sup>** | **Sustainment**<br>**N = 24,498<sup>I</sup>** | **Maintenance**<br>**N = 8,246<sup>I</sup>** |
| --- | --- | --- | --- | --- |
| **Age (years)** | 10 (5, 15) | 10 (5, 15) | 10 (5, 15) | 10 (5, 15) |
| **Sex** |  |  |  |  |
| **Female** | 18,574<br>(52%) | 1,516 (51%) | 12,750<br>(52%) | 4,308 (52%) |
| **Male** | 17,122<br>(48%) | 1,440 (49%) | 11,748<br>(48%) | 3,934 (48%) |
| **X (non-binary)** | 4 (<0.1%) | 0 (0%) | 0 (0%) | 4 (<0.1%) |
| **Race & Ethnicity** |  |  |  |  |
| **American Indian or Alaska Native** | 188 (0.5%) | 15 (0.5%) | 119 (0.5%) | 54 (0.7%) |
| **Asian** | 2,336 (6.5%) | 182 (6.2%) | 1,602 (6.5%) | 552 (6.7%) |
| **Black or African American** | 1,516 (4.2%) | 131 (4.4%) | 1,032 (4.2%) | 353 (4.3%) |
| **Hispanic** | 7,166 (20%) | 615 (21%) | 4,861 (20%) | 1,690 (20%) |
| **Multiracial** | 1,782 (5.0%) | 165 (5.6%) | 1,179 (4.8%) | 438 (5.3%) |
| **Native Hawaiian or Other Pacific Islander** | 77 (0.2%) | 9 (0.3%) | 53 (0.2%) | 15 (0.2%) |
| **Non-Hispanic White** | 19,438<br>(54%) | 1,572 (53%) | 13,514<br>(55%) | 4,352 (53%) |
| **Other** | 1,447 (4.1%) | 109 (3.7%) | 981 (4.0%) | 357 (4.3%) |
| **Unknown/Declined** | 1,750 (4.9%) | 158 (5.3%) | 1,157 (4.7%) | 435 (5.3%) |
| **PMCA Category** |  |  |  |  |
| **Complex Chronic** | 12,303<br>(34%) | 1,179 (40%) | 8,778 (36%) | 2,346 (28%) |
| **Non-chronic** | 6,030 (17%) | 455 (15%) | 3,937 (16%) | 1,638 (20%) |
| **Non-complex Chronic** | 6,058 (17%) | 522 (18%) | 4,194 (17%) | 1,342 (16%) |
| **Not Categorized** | 11,309<br>(32%) | 800 (27%) | 7,589 (31%) | 2,920 (35%) |
| **Insurance Type** |  |  |  |  |
| **Commercial Insurance** | 19,943<br>(56%) | 1,583 (54%) | 13,605<br>(56%) | 4,755 (58%) |
| **Government Insurance** | 15,757<br>(44%) | 1,373 (46%) | 10,893<br>(44%) | 3,491 (42%) |
| **Language Group** |  |  |  |  |
| **English** | 32,501<br>(91%) | 2,662 (90%) | 22,260<br>(91%) | 7,579 (92%) |
| **Language Other than English** | 3,199 (9.0%) | 294 (9.9%) | 2,238 (9.1%) | 667 (8.1%) |
| **Home Location** |  |  |  |  |
| **Seattle/Tacoma/Everett** | 21,597<br>(60%) | 1,717 (58%) | 14,705<br>(60%) | 5,175 (63%) |
| **Ferry, far** | 655 (1.8%) | 44 (1.5%) | 462 (1.9%) | 149 (1.8%) |
| **Ferry, near** | 1,462 (4.1%) | 109 (3.7%) | 1,015 (4.1%) | 338 (4.1%) |
| **Out of State/Unknown** | 2,436 (6.8%) | 248 (8.4%) | 1,724 (7.0%) | 464 (5.6%) |
| **Pass, far** | 1,339 (3.8%) | 104 (3.5%) | 889 (3.6%) | 346 (4.2%) |
| **Pass, near** | 4,529 (13%) | 405 (14%) | 3,206 (13%) | 918 (11%) |
| **Western WA, far** | 3,682 (10%) | 329 (11%) | 2,497 (10%) | 856 (10%) |

| **Appointment Type** |  |  |  |  |
| --- | --- | --- | --- | --- |
| **In-person Appointment** | 17,821<br>(50%) | 773 (26%) | 11,807<br>(48%) | 5,241 (64%) |
| **Telehealth Appointment** | 16,612<br>(47%) | 1,223 (41%) | 12,392<br>(51%) | 2,997 (36%) |
| **Telephone Appointment** | 1,267 (3.5%) | 960 (32%) | 299 (1.2%) | 8 (<0.1%) |
| <sup>†</sup> Median (Q1, Q3); n (%) |  |  |  |  |

Telephone appointments were scheduled as in-person visits and converted due to COVID; for modeling purposes, telephone visits are counted as telehealth.
PMCA – Pediatric Medical Complexity Algorithm

The trend of appointment types from prior to 2020 to 2024 are depicted in **Figure 1** by visit count and percent of total utilization per month. Telephone appointments were used most frequently in the initial phase of the pandemic. Telemedicine usage notably decreased after 2021, with stable levels throughout 2022-2024.

**Figure 1.**
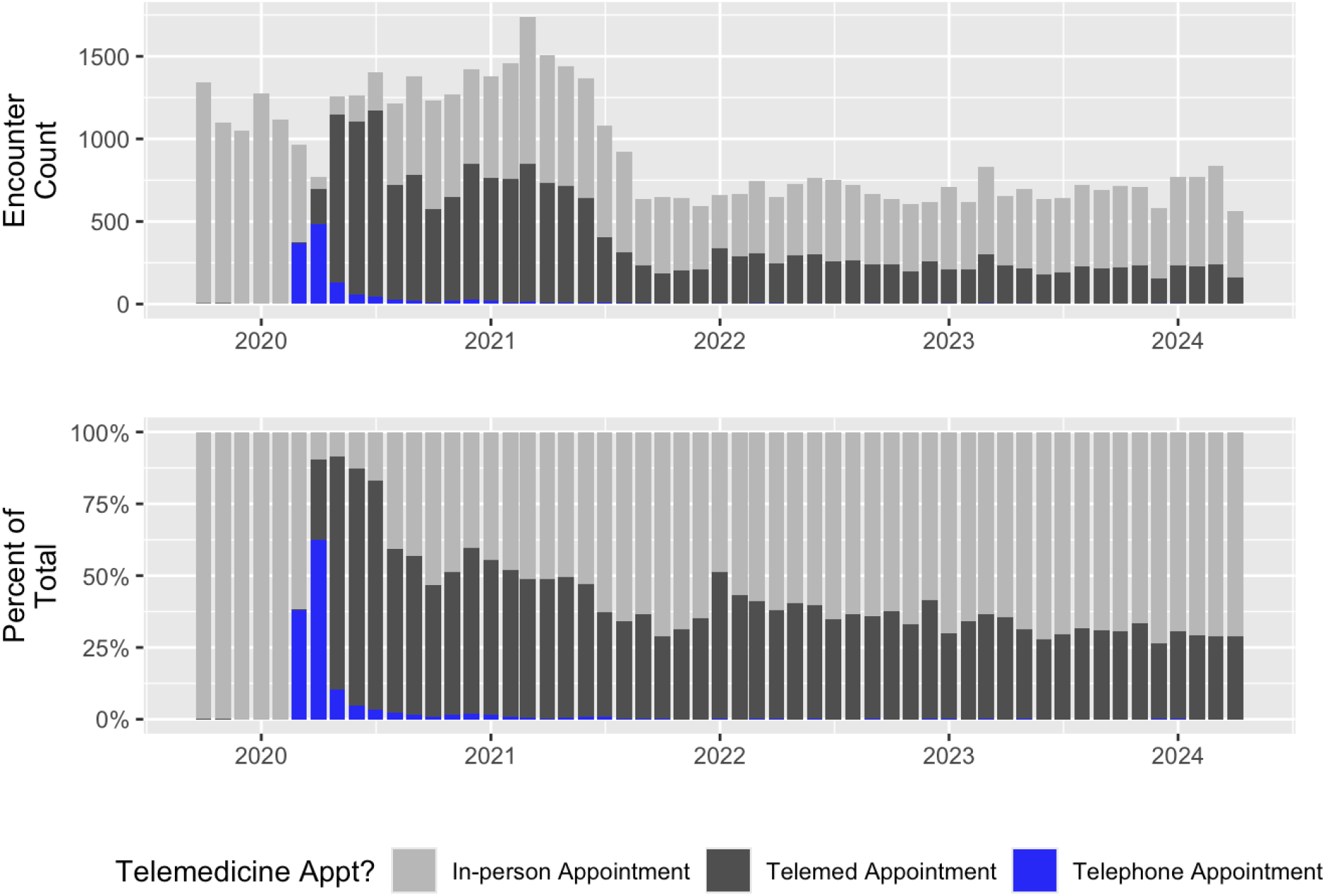
Telehealth encounter count and percent of total utilization by month.

The odds of telehealth utilization across demographic groups during the three pandemic phases is described in **Table 2**. Asian patients had decreased usage in the maintenance phase [OR 0.62 (0.48, 0.81)]. Black or African American patients demonstrated consistently lower telehealth utilization across all phases, with statistically significant reduced odds during the Sustainment phase (OR 0.86, 95% CI 0.73-1.00). Non-Hispanic White patients exhibited progressively increasing telehealth utilization, with the highest odds during the Maintenance phase (OR 1.41, 95% CI 1.22-1.61). Non-English speakers had significantly decreased telehealth utilization during the sustainment and maintenance phases of the pandemic (OR 0.56, 95% CI 0.49, 0.64); (OR 0.50, 95% CI 0.37-0.68) compared to English speakers.

**Table 2.** Adjusted odds ratios of telehealth utilization during and after the COVID-19 pandemic.

| Variable<sup>1</sup> | Initial |  | Sustainment |  | Maintenance |  |
| --- | --- | --- | --- | --- | --- | --- |
|  | OR | 95% CI | OR | 95% CI | OR | 95% CI |
| **Race & Ethnicity** |  |  |  |  |  |  |
| Asian | 1.43 | 1.00, 2.04 | 0.98 | 0.86, 1.11 | **0.62** | 0.48, 0.81 |
| Black or African American | 0.74 | 0.51, 1.10 | **0.86** | 0.73, 1.00 | 0.79 | 0.58, 1.07 |
| Hispanic | 1.08 | 0.85, 1.37 | 0.95 | 0.87, 1.04 | 1.04 | 0.86, 1.25 |
| Non-Hispanic White | 1.14 | 0.95, 1.37 | **1.15** | 1.08, 1.23 | **1.41** | 1.22, 1.61 |
| Other | 0.79 | 0.60, 1.04 | 1.09 | 0.98, 1.21 | **1.28** | 1.04, 1.58 |
| Unknown/Declined | 0.96 | 0.59, 1.56 | 1.01 | 0.84, 1.21 | 1.09 | 0.75, 1.58 |
| **Language of Care** |  |  |  |  |  |  |
| English | 1.00 | — | 1.00 | — | 1.00 | — |
| Language Other than English | 0.74 | 0.52, 1.05 | **0.56** | 0.49, 0.64 | **0.50** | 0.37, 0.68 |
| **Payor Mix** |  |  |  |  |  |  |
| Commercial Insurance | 1.00 | — | 1.00 | — | 1.00 | — |
| Government Insurance | 1.17 | 0.94, 1.44 | **0.91** | 0.84, 0.98 | **0.83** | 0.71, 0.98 |
| **PMCA Category** |  |  |  |  |  |  |
| Complex Chronic | 1.00 | — | 1.00 | — | 1.00 | — |
| Non-chronic | 0.91 | 0.68, 1.23 | **0.67** | 0.60, 0.75 | **0.45** | 0.36, 0.56 |
| Non-complex Chronic | 0.98 | 0.74, 1.29 | **0.81** | 0.73, 0.91 | **0.59** | 0.47, 0.75 |
| Not Categorized | 0.79 | 0.62, 1.00 | **0.61** | 0.56, 0.67 | **0.36** | 0.30, 0.44 |
| **Home Location** |  |  |  |  |  |  |
| Seattle/Tacoma/Everett | 1.00 | — | 1.00 | — | 1.00 | — |
| Ferry, far | 0.93 | 0.42, 2.03 | **1.67** | 1.27, 2.21 | **1.96** | 1.12, 3.42 |
| Ferry, near | 1.11 | 0.66, 1.86 | **1.58** | 1.32, 1.89 | **1.51** | 1.05, 2.17 |
| Out of State/Unknown | 1.45 | 0.98, 2.12 | **1.93** | 1.66, 2.24 | **2.59** | 1.86, 3.60 |
| Pass, far | 1.16 | 0.67, 1.98 | **2.26** | 1.85, 2.76 | **4.10** | 2.77, 6.07 |
| Pass, near | 1.35 | 0.99, 1.85 | **2.38** | 2.12, 2.68 | **1.76** | 1.38, 2.25 |
| Western WA, far | 1.10 | 0.79, 1.52 | **1.34** | 1.18, 1.52 | 1.15 | 0.89, 1.48 |

<sup>1</sup> Deviation contrasts are used for the Race & Ethnicity variable; all other variables are assessed using treatment contrasts

Patients with government insurance (i.e. Medicaid) had decreased usage during the sustainment and maintenance phases compared to those with commercial insurance (OR 0.91, 95% CI 0.84-0.98; OR 0.83, 95% CI 0.71-0.98). Patients with complex chronic conditions utilized telehealth more frequently than those with non-chronic conditions, whose telehealth utilization comparatively decreased from the pandemic to the post-pandemic phases (OR 0.91 to 0.45). Geographic location was strongly associated with telehealth utilization, with patients living farther from the medical center showing significantly higher odds of telehealth use. Patients in the “ Pass, far” rural region (across the Cascade mountains with >90 minutes travel from tertiary care site) had the highest telehealth utilization during the Maintenance phase (OR 4.10, 95% CI 2.77-6.07), and those in “ Out of State/Unknown” locations also showed consistently high telehealth use (OR 2.59, 95% CI 1.86-3.60 during Maintenance).

The probability of telehealth usage across the three pandemic phases stratified by race/ethnicity, insurance type, and language preference is depicted in **Figure 2**. Across all phases, non-English speakers consistently demonstrated lower telehealth utilization compared to their English-speaking counterparts within the same racial/ethnic and insurance groups, with this disparity widening during the Maintenance Phase.

**Figure 2.**
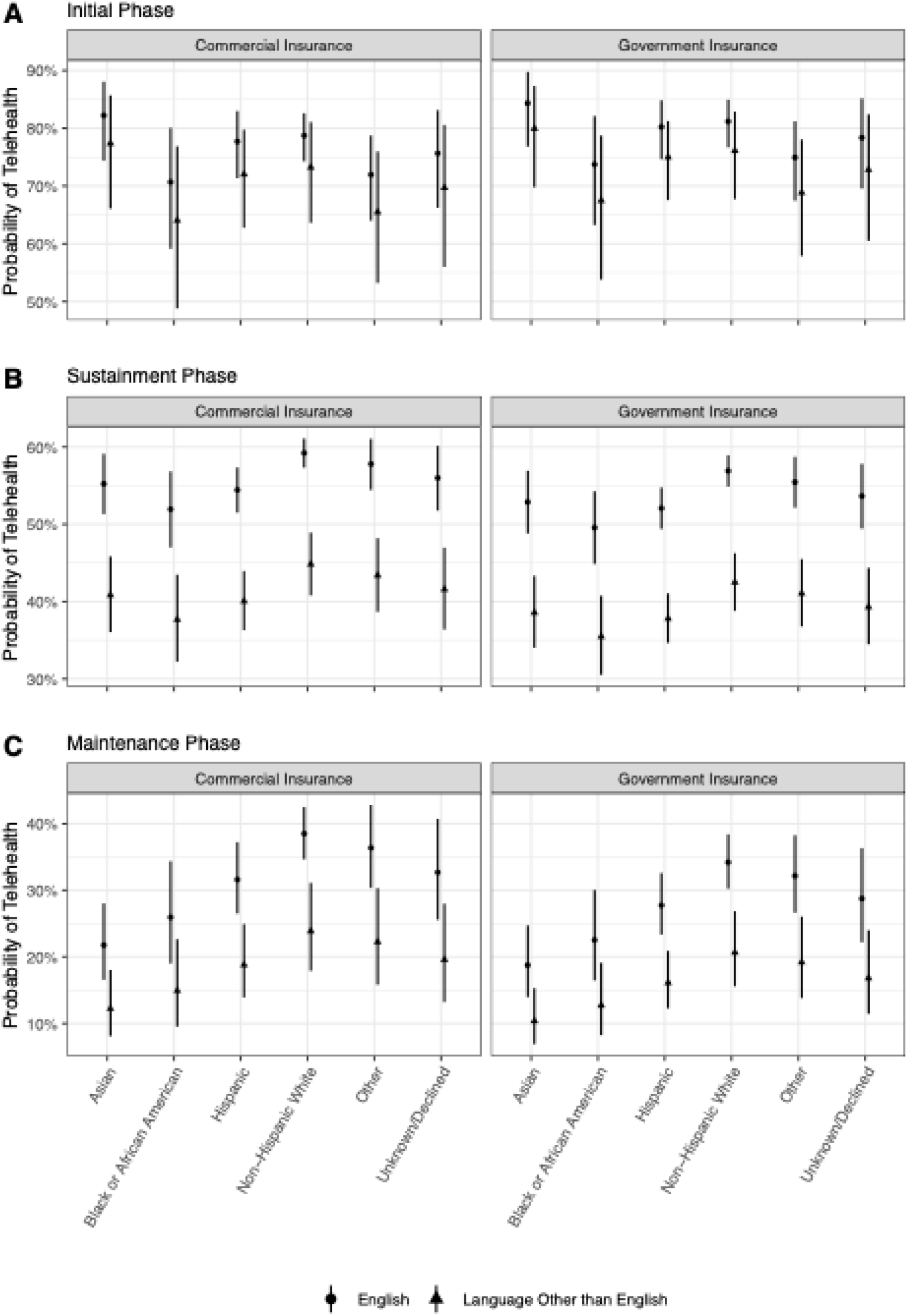
Regression effect sizes by race/ethnicity and type of insurance across telehealth implementation phases. A. Initial: March 2020-May 2020 B. Sustainment: June 2020-March 2022 C. Maintenance: April 2022-March 2023

## Discussion

The COVID-19 pandemic catalyzed rapid adoption of telehealth throughout the healthcare system, creating an opportunity to examine utilization patterns across various demographic groups and geographic regions.^12,13^ Understanding telehealth utilization patterns is essential to understanding what populations are benefitting most from this care modality. It can also help inform policies and targeted interventions to increase telehealth accessibility and usage. This study found that rural populations had increased and sustained usage of telehealth even post-pandemic. It also found that a non-English primary language, those on government insurance, and being Black/African American were predictors for decreased telehealth usage.

Throughout the pandemic, various improvements were made to the telehealth clinical workflow at Seattle Children’s Hospital, such as improved telehealth interpreter access and simplification of appointment notification and access to Zoom, all aimed to improve access to telehealth care. Despite these changes, the long-term impact of the pandemic on telehealth usage remains mixed.^14^ This retrospective analysis of telehealth utilization during and after the pandemic within a pediatric neurology department that services a large, geographically diverse region identified demographic differences in utilization patterns across time and highlights the importance of preserving telehealth access especially for rural communities.

### Cohort Representativeness in Washington State

This study’s population closely mirrors the demographic makeup of Washington state’s pediatric population.^15^ The racial and ethnic distribution of this cohort – where non-Hispanic White patients make up the largest group, followed by significant representation from Black, Asian, Hispanic, and other racial/ethnic groups – aligns with statewide population estimates for children and adolescents. Insurance coverage in this cohort, with a majority of patients covered by commercial insurance, also reflects patterns seen across Washington state’s pediatric population. Accordingly, these findings are likely generalizable to the broader Washington state population, and similar telehealth utilization patterns would be anticipated if this analysis were conducted in another representative sample from across the state.

### Evolution of Telehealth Utilization Patterns

The decline in telehealth usage as the pandemic subsided is an expected trend and aligns with the natural evolution of healthcare delivery during this time period. In the initial phase, telephone appointments were most frequently utilized, reflecting the lack of widespread integration and familiarity with more robust video-based telehealth platforms. This is reflected in other studies, which document the rapid transition to telehealth in response to the pandemic, with an initial reliance on more simple technology followed by the gradual adoption of more sophisticated platforms.^16,17^

### Geographic Differences in Telehealth Utilization

Geographic location emerged as a strong predictor for telehealth utilization, with patients that live farther from the city demonstrating significantly higher odds of telehealth usage throughout all phases. This persistent pattern emphasizes the critical role telehealth plays in eliminating barriers related to travel distance and time away from school and work, particularly for families in more rural areas.^18^ The sustained high utilization among geographically distant patients in the maintenance phase suggests telehealth has become an established norm for these populations, and highlights the importance of maintaining telehealth access for these communities post-pandemic to provide care that might otherwise not be accessible.^19^ Sustaining telehealth access with payment parity with in-person visits, expanded insurance coverage, and continued flexibility in licensing requirement would support a long-term sustainability of telehealth care delivery. Furthermore, the utilization of telehealth has also been associated with lower high-cost hospital utilization and could potentially be a cost-saving measure for the healthcare system overall.^20^

### Demographic Differences in Utilization Patterns

This study also identified differences in telehealth utilization across various demographic factors. These differences suggest areas of opportunities to enhance telehealth services for certain populations. Since utilization patterns are likely influenced by a variety of factors including differences in social determinants and historical influences, interpreting the cause of changes and gaps in utilization patterns is limited without survey or qualitative data from each population.

Black or African American patients had consistently lower telehealth utilization throughout all pandemic phases, which aligns with previous studies that have shown significantly lower telehealth adoption in this population.^4,21,22^ The persistence of this difference even post-pandemic could be attributed to broader trends of healthcare underutilization and untrustworthiness overall within this population; prior studies have also suggested that structural barriers and systemic inequities contribute to this divide.^23,24^ Asian patients also showed decreased telehealth utilization that became statistically significant in the maintenance, suggesting an emerging change in utilization that warrants close monitoring in the post-pandemic period. Further data regarding barriers, resources and preferences in these groups is needed to contextualize this pattern and begin to develop potential interventions.

Patients with complex chronic conditions were significantly more likely to utilize telehealth across all three pandemic phases, with a wider gap in the maintenance phase. This could reflect their need for frequent specialist follow-up and care coordination that is conducive for telehealth. For pediatric neurology patients, who often face mobility issues, telehealth offers critical benefits such as minimizing travel burdens, maintaining continuity of care, and enabling timely intervention for acute issues. These advantages are particularly important for families managing complex multiorgan system disorders, as telehealth can facilitate multidisciplinary collaboration. As a result, telehealth remains an essential modality for ensuring access and quality care for children with complex medical needs.

Non-English speakers demonstrated significantly decreased utilization during both sustainment and maintenance phases compared to English speakers, even after adjusting for confounding demographic factors. Language emerged as a powerful determinant of telehealth utilization, creating a compounded effect for decreased telehealth access among non-English speakers across all racial/ethnic groups. This finding illustrates the intersectional nature of telehealth disparities and suggests that targeted interventions may need to address multiple barriers simultaneously to substantially improve access and utilization.

Limited availability of multilingual educational materials about telehealth services, insufficient interpreter access prior to virtual visits to assist with joining, and the possibility that telehealth may not be consistently offered as an option to non-English speaking families as a follow-up option have all been previously identified areas of improvement.^25–27^ Increasing virtual interpreter resource availability, proactively offering telehealth visits to non-English speakers, and improving language concordant patient education resources for telehealth may all be possible solutions for improved access to telehealth care.

### Limitations

This study has several limitations. The analysis relied on cross-sectional data from a single large healthcare system, which limits its generalization to other geographic regions or healthcare settings with different telehealth infrastructure and patient populations. In addition, more detailed socioeconomic factors such as digital literacy levels and technology availability that likely play a role in telehealth utilization patterns were not assessed through this retrospective study. Data regarding internet access at home was not available, which is seen a common target for bridging the telehealth divide. Although several changes were implemented throughout the pandemic to improve telehealth access, the specific dates of implementation and details behind these changes were unavailable for this study. Similarly, rapidly evolving telehealth policies during the pandemic makes it difficult to isolate the effects of our variables from concurrent policy changes. Future research could address these limitations through mixed methods approaches that incorporate qualitative data on policy implementation patterns and patient experiences and preferences.

## Conclusions

This study’s findings underscore the importance of maintaining telehealth as an option for all patients, especially for those with complex medical needs and those residing in rural areas who demonstrated sustained high utilization throughout all pandemic phases. The continued relevance of telehealth for these populations exemplifies its potential to address longstanding geographic disparities in access to specialized pediatric care.

This study also identified multiple persistent differences in utilization, particularly for non-English speakers, that have important implications for telehealth implementation in pediatric specialty care. As telehealth has become a more established component of healthcare delivery, ensuring equitable access will require targeted interventions addressing specific barriers faced by underserved populations.

Strategies could include expanding interpreter services for telehealth visits within the hospital system, creating multilingual educational materials about telehealth options to be given to patients at in-person visits, addressing digital literacy gaps, and implementing outreach programs for communities with lower telehealth utilization.

## Data Availability

All data produced in the present study are available upon reasonable request to the authors.

## Declarations

### Ethical approval

Seattle Children’s IRB determined this study to be exempt given that the goal of this study is for quality improvement in care delivery and that the patient data used were deidentified at onset.

### Consent

Not applicable, deidentified dataset.

### Availability of data and materials

The datasets generated and/or analyzed during the current study are not publicly available but are potentially available from the corresponding author on reasonable request.

### Disclaimers

None

### Author contributions

PC: Conceptualization, Methodology, Investigation, Writing – Original Draft, Writing – Review & Editing, Project administration

DB: Methodology, Software, Validation, Formal analysis, Visualization, Writing – Review & Editing

LM: Writing – Review & Editing

MW: Writing – Review & Editing

PP: Conceptualization, Methodology, Investigation, Writing – Review & Editing, Project administration

